# The Influence of Gender on Diagnostic Markers of Acute Kidney Injury in Acute-on-Chronic Liver Failure

**DOI:** 10.1101/2025.04.23.25326257

**Authors:** Rohini Saha, Subhadra Priyadarshini, Shalimar, Pragyan Acharya

## Abstract

Gender has a profound impact on disease severity, outcomes and diagnosis yet, its role in clinical disease is insufficiently explored. Acute on chronic liver failure (ACLF) is associated with high mortality and multiple organ dysfunction, where acute kidney injury (AKI) significantly worsens prognosis. Here we investigated the impact of gender on the diagnostic parameters used for severity grading in ACLF. We enrolled 1134 ACLF patients, and shortlisted 757 patients (636 males, 121 females) admitted to AIIMS, New Delhi, between 2016 and 2023. ACLF-AKI was defined and staged according to International Club of Ascites (ICA) criteria. The impact of gender on baseline clinical parameters, AKI incidence, and progression were assessed using the statistical tools IBM SPSS 26.0 and GraphPad Prism Males exhibited a higher incidence of AKI (48.34%) compared to females (28.09%). However, no significant gender-based differences were observed in AKI stages. Males also had an overall high absolute value of sCr and blood urea compared to females. However, female ACLF patients who developed AKI exhibited a significantly higher δ sCr levels compared to males (p=0.003). In conclusion, gender-based differences were observed in the widely used diagnostic criteria of sCr and δ sCr for AKI in patients with ACLF. Although these findings are preliminary our results reveal gender-specific differences in sCr-based AKI diagnosis and risk stratification in ACLF. These results inform the need for deeper exploration of gender influenced parameters in ACLF specifically in AKI diagnosis.

## Introduction

Gender is a key determinant of human physiology, influencing the expression of numerous genes and proteins through the presence of XX or XY sex chromosomes. While certain diseases exhibit clear gender biases in incidence and outcomes, the influence of gender on diagnostic parameters remains incompletely understood.

Acute-on-chronic liver failure (ACLF) is a severe complication of cirrhosis characterized by high short-term mortality and multi-organ dysfunction (1,2). Among these complications, acute kidney injury (AKI) markedly increases the risk of mortality (3, 4). Currently, ACLF management is largely supportive, with no targeted therapies available due to limited understanding of the disease’s underlying mechanisms (5, 6). An effective strategy for improving outcomes in ACLF would be identification of patients at risk for AKI and the implementation of nephroprotective interventions at an early stage (7).

Serum creatinine (sCr) is the primary biomarker used to diagnose AKI across etiologies, including ACLF. However, sCr levels are affected by various factors, such as diet, muscle mass, and hydration status (8). In patients with cirrhosis, including those with ACLF, the International Club of Ascites (ICA) defines AKI based on an increase in sCr from baseline (δ sCr), which is an improved diagnostic criteria (9). However, the role of gender in determining AKI diagnostic thresholds in ACLF remains poorly characterized.

In this study, we investigate gender-based differences in AKI outcomes and diagnostic parameters in a well-characterized cohort of ACLF patients (n = 757).

## Methods

### Study participants

We recruited a total of 1134 patients, admitted with ACLF diagnosis, to the department of gastroenterology, AIIMS, New Delhi, during the period of 2016-2023. The inclusion criteria included ACLF patients defined by the EASL criteria for grades of organ failure. Patients with Grade 0 were excluded. The other exclusion criteria included presence of HCC, diabetes, use of ATT-drugs or herbal medicines, or patients without clinical medical record. The gender of the patients were assigned based on biological parameters, that is males (n=636) and females (n=121).

We used the International Club of Ascites (ICA) (9) criteria to define acute kidney injury (AKI) and no-AKI and also assign stages and type of AKI. This criterion defines AKI as an increase in sCr ≥0.3 mg/dL (≥26.5 mmol/L) from baseline within 48 hour or a percentage increase in sCr ≥50% from baseline. The stages of AKI were assigned as:

- Stage 1: increase in sCr ≥0.3 mg/dl (26.5 μmol/L) or an increase in sCr ≥1.5-fold to 2-fold from baseline
- Stage 2: increase in sCr >2-fold to 3-fold from baseline
- Stage 3: increase of sCr >3-fold from baseline or sCr ≥4.0 mg/dl (353.6 μmol/L) with an acute increase ≥0.3 mg/dl (26.5 μmol/L) or if the patient requires a renal replacement therapy (RRT).

For assigning progression to AKI, patients were followed up during the hospital stay and AKI development was assessed following the ICA criteria. The delta sCr (δsCr) was calculated as the difference between the creatinine levels on day 0 and the highest creatinine levels on subsequent days (3, 7, or 14 days).

### Statistical analysis and data representation

Continuous data were presented as median with inter quartile range (Q1-Q3), while categorical data presented as frequency and percentage. Initially, bivariate analysis was performed to assess associations, using Mann-Whitney U-test for continuous variables and Chi-square/Fisher’s exact test for categorical variables. Univariate and multivariate logistic regression was conducted to identify independent predictors of acute kidney injury (AKI). The results are reported as odds ratios (OR) with 95% confidence intervals (CI). Variables with potential significance (p < 0.20) in the univariate regression analysis were then included in the multivariate model to adjust for confounders. Multicollinearity among variables was checked before proceeding with the multivariate regression analysis. Statistical significance was set at p < 0.05. For data visualization, Box-whiskers plots were plotted for variables with continuous data points. The median was represented with a horizontal line within each of the box, and the whiskers represent the maximum and minimum data point (11).

Vertical histogram plots were used to plot the percentages or fractions of the total population of patients, derived from a Chi-square contingency table. The contingency table organizes data by placing one data set in rows and another in columns. The cells where the rows and columns intersect show the percentage of cases for each combination of categories (12,13), for instance AKI incidence and AKI stages. These are single-observations, hence no standard deviation or error bars were plotted. All analyses and data visualization were conducted using statistical software IBM SPSS version 26.0 and GraphPad Prism version 8.

## Results

In the overall ACLF cohort, baseline levels of serum urea and creatinine were significantly higher in males compared to age- and ACLF grade-matched females (urea: 76 mg/mL vs. 51 mg/mL; creatinine: 1.9 mg/dL vs. 1.4 mg/dL; *p* ≤ 0.0001 for both; Figure 1A, 1B). To further evaluate these differences, patients were stratified by AKI status and stage according to ICA criteria (9).

**Figure 1.**
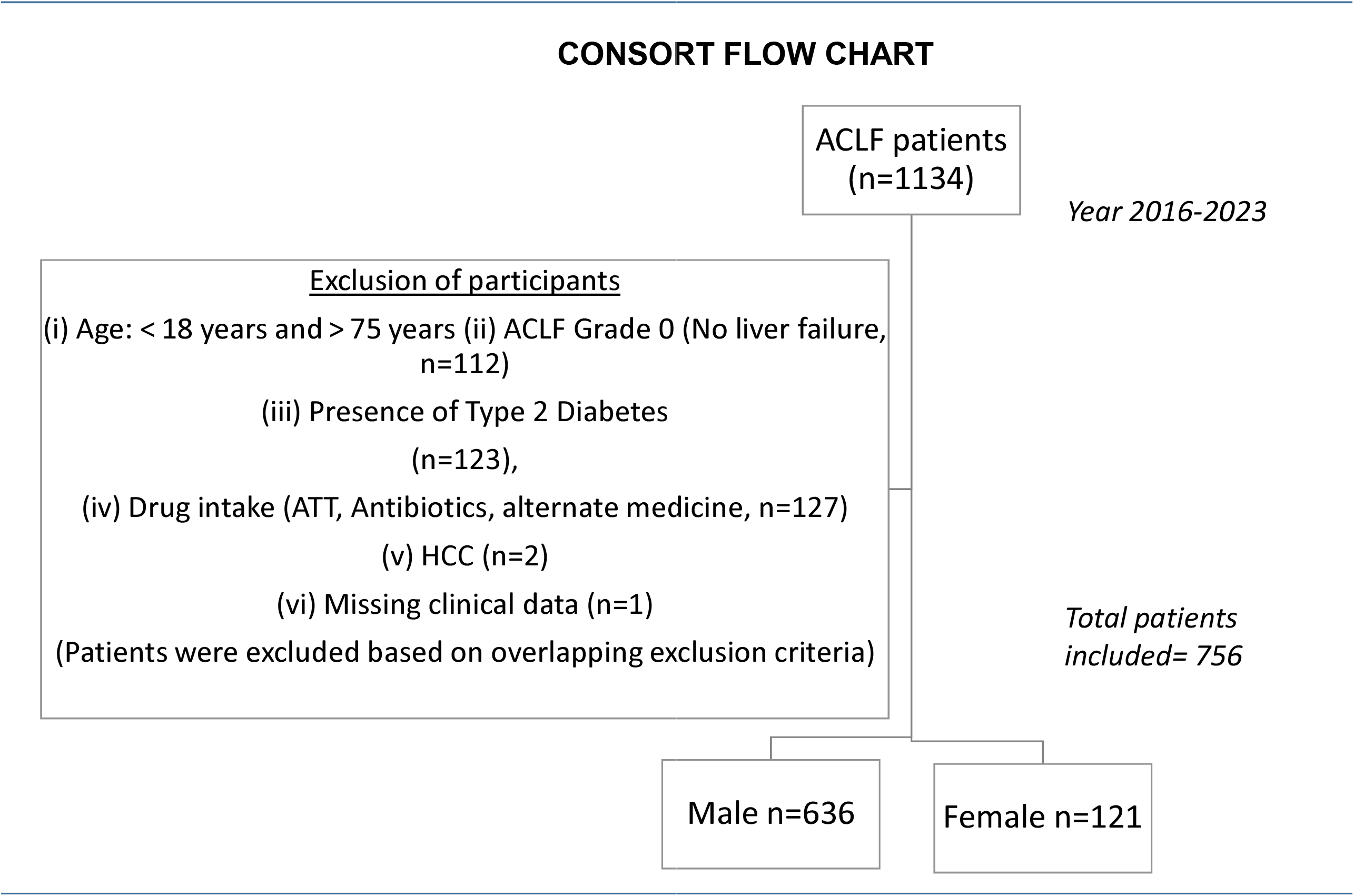
CONSORT Flow Chart. A total of n=1134 study participants were recruited in the study. Based on the exclusion criteria (see Supplemental Methods), n=378 patients were excluded from the study. Among the included patients (n=757), 83.9% patients were males (n=636) and 16% patients were females (n=121)

**Figure 2.**
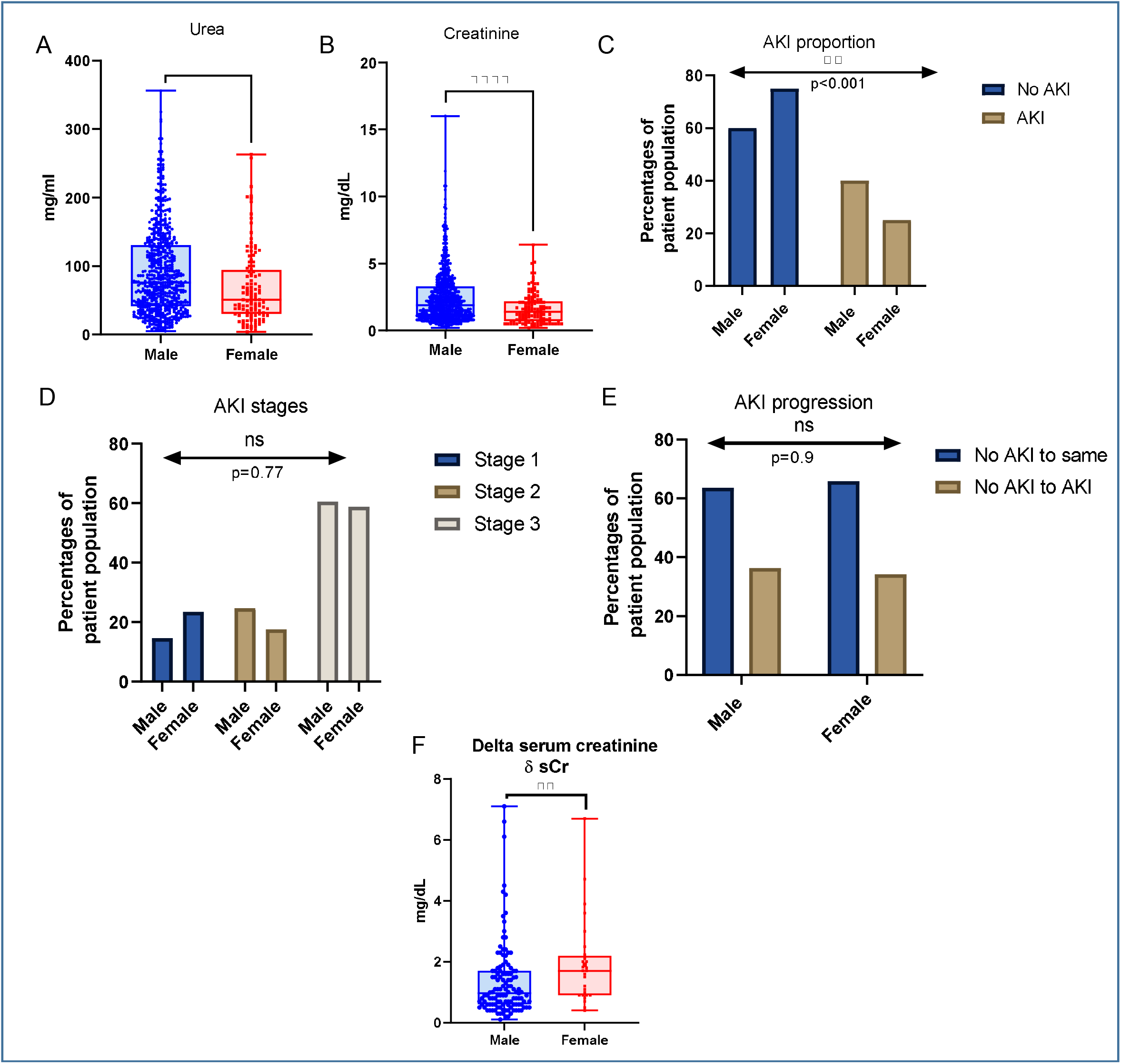
Gender influences AKI in ACLF patients. A. Levels of blood urea (mg/mL) are significantly different in males versus females, p<0.0001; B. Levels of sCr (mg/dL) are higher in males vs females, p<0.0001, C. Males have a higher incidence of AKI than females, p<0.001) D. The distribution of stages AKI (1, 2, 3) is not influenced by gender (p=0.77) E. The percentage of ACLF no-AKI patients who progress to AKI (no-AKI to AKI) or those who do not progress (no-AKI to same) have equivalent proportion of males and females (p=0.937), F. Females (n=28) who progress from no-AKI to AKI show a higher rise in sCr levels as compared to males (n=123) who progress to AKI (expressed as delta serum creatinine, p=0.003). A, B, and F represent box-plots with median central tendency line within each box. Each dots represents individual patient values. C-E represents vertical bar plots depicts the percentage of total males or female in the cohort, derived from a Chi-squared contingency table. These percentages of patient population are single observations; hence S.D. or error bars are not plotted (See Methods)

A higher proportion of male patients developed AKI (48.34%) compared to females (28.09%) (Figure 1C, Table 1), though no significant differences were observed in AKI staging between genders (Figure 1D, Table 1). Similarly, the proportion of patients who progressed to AKI during hospitalization (within 14 days) was comparable between sexes (Figure 1E, Table 1). Among patients who progressed to AKI, the magnitude of change in serum creatinine (δ sCr) was significantly greater in females (median δ sCr = 1.7 mg/dL) compared to males (median δ sCr = 0.97 mg/dL; *p* = 0.003) (Figure 1F, Table 1).

**Table 1.** Baseline parameters in ACLF males (n=636) and females (n=121) included in the study. Continuous data are presented as median with inter quartile range (Q1-Q3), while categorical data presented as percentage. Bivariate analysis is performed to determine p-value significance, using Mann-Whitney U-test for continuous variables and Chi-square/Fisher’s exact test for categorical variables.

| **Parameters** | **Female (n=121)** | **Male (n=636)** | **p-value** |
| --- | --- | --- | --- |
| Age (years) | 40 (30-50) | 40 (35-48) | 0.683 |
| Haemoglobin (g/dL) | 8.5 (7.2-10.4) | 8.4 (6.8-10.0) | 0.232 |
| TLC (per mm<sup>3</sup>) | 9860 (5850-16135) | 13300 (8200-20000) | <0.001 |
| Platelets (x10<sup>3</sup> per mm<sup>3</sup>) | 89.5 (59.0-142.0) | 92.0 (60.0-139.0) | 0.939 |
| INR | 2.700 (1.870-3.740) | 2.600 (2.060-3.500) | 0.771 |
| Urea (mg/mL) | 51.00 (30.00-93.50) | 76.00 (41.00-130.50) | <0.0001 |
| Creatinine (mg/dL) | 1.40 (0.70-2.15) | 1.90 (1.10-3.30) | <0.0001 |
| Sodium (mEq/L) | 136.0 (132.0-140.0) | 136.0 (131.0-141.0) | 0.358 |
| Potassium (mEq/L) | 3.90 (3.36-4.65) | 4.20 (3.70-5.00) | 0.003 |
| Bilirubin (mg/dL) | 15.15 (4.75-22.70) | 12.70 (5.75-22.70) | 0.997 |
| Aspartate<br>Aminotransferase (AST)<br>(IU/L) | 115.0 (58.0-216.0) | 113.0 (68.0-182.0) | 0.861 |
| Alanine Aminotransferase<br>(ALT) (IU/L) | 56 (30-117) | 45 (31-77) | 0.181 |
| Alkaline Phosphatase<br>(ALP) (IU/L) | 236.0 (154.0-347.0) | 183.5 (130.0-271.0) | 0.002 |
| Albumin (g/dL) | 2.50 (2.10-2.90) | 2.50 (2.20-2.90) | 0.581 |
| Globulin (g/dL) | 3.00 (2.50-3.70) | 3.40 (2.70-4.10) | 0.011 |
| Presence of Infection (%) | 63 (53.4%) | 330 (52.9%) | 0.920 |
| AKI Proportion (%) | 34 (28.09%) | 307 (48.34%) | <0.001 |
| Progression to AKI (%) | 28 (34.15%) | 123 (36.39%) | 0.937 |
| $\delta$ sCr (mg/dL) | 1.7 (0.9-2.20) | 0.97 (0.6-1.7) | 0.003 |
| AKI Stage 1 | 8 (23.5%) | 45 (14.65%) | 0.77 |
| AKI Stage 2 | 6 (17.64%) | 76 (24.75%) |  |
| AKI Stage 3 | 20 (58.82%) | 186 (60.58%) |  |
| Alcohol consumer (%) | 8 (6.8%) | 494 (79.9%) | <0.001 |
| 28-day Mortality rate (%) | 75 (62.0%) | 446 (70.2%) | 0.072 |
| ACLF Grade 1 | 12 (11.2%) | 42 (7.3%) | 0.524 |
| ACLF Grade 2 | 27 (25.2%) | 147 (25.6%) |  |
| ACLF Grade 3 | 68 (63.6%) | 385 (66.6%) |  |

These findings suggest that although fewer females developed ACLF-AKI, those who did experienced a more rapid and pronounced increase in serum creatinine. This may indicate a narrower window for early AKI detection and intervention in female patients, emphasizing the need for heightened clinical vigilance and potentially gender-specific monitoring strategies in ACLF.

Univariate analysis identified serum creatinine as the strongest predictor of AKI in both genders, with an odds ratio (OR) of 9.018 in males and 14.037 in females (Table 2). In males, additional predictors included ACLF grade, presence of infection, alcohol use, serum urea, and sodium. Notably, serum globulin appeared to have a protective effect against AKI in males (OR = 0.676; Table 2). In contrast, for females, other significant variables influencing AKI included potassium, urea, and sodium (Table 2). These observations suggest gender-based differences in the underlying pathophysiology of AKI in ACLF where systemic factors may play a larger role in males, while renal-specific parameters are more predictive in females.

**Table 2.** Univariate logistic regression analysis of factors affecting presence of AKI among males and females. The results are reported as odds ratios (OR) with 95% confidence intervals (CI).

| 1. Male | No AKI | AKI | Univariate Logistic Regression |  |  |
| --- | --- | --- | --- | --- | --- |
|  |  |  | p-value | Odds ratio (OR) | 95% CI of OR |
| Infection (Yes) | 135 (45.6%) | 169 (59.7%) | <□.001 | 1.768 | (1.271-2.459) |
| Alcohol (Yes) | 225 (76.5%) | 231 (83.4%) | 0.042 | 1.540 | (1.016-2.334) |
| ACLF Grade at admission |  |  |  |  |  |
| 1 | 26 (9.9%) | 9 (3.4%) | *Reference category* |  |  |
| 2 | 82 (31.2%) | 45 (17.1%) | 0.283 | 1.585 | (0.684-3.675) |
| 3 | 155 (58.9%) | 209 (79.5%) | <□0.001 | 3.895 | (1.775-8.548) |
| Age (years) | 40 (35-47) | 40 (35-49) | 0.245 | 1.00 | (0.994-1.030) |
|  |  |  |  | 9 |  |
| Hemoglobin (g/dL) | 8.5 (6.9-10.3) | 8.2 (6.6-9.8) | 0.039 | 0.933 | (0.874-0.997) |
| TLC (per mm<sup>3</sup>) | 11500 (7700-18000) | 15250 (8950-23565) | <□0.001 | 1.000 | (1.000-1.000) |
| Platelets (x10<sup>3</sup> per mm<sup>3</sup>) | 90.0 (61.0-141.0) | 92.0 (58.5-130.5) | 0.372 | 1.001 | (0.999-1.000) |
| INR | 2.590 (2.045-3.310) | 2.700 (2.070-3.600) | 0.173 | 1.060 | (0.975-1.150) |
| Urea (mg/mL) | 47.00 (28.00-77.00) | 123.00 (76.00-167.00) | <□0.001 | 1.027 | (1.022-1.032) |
| Creatinine (mg/dL) | 1.10 (0.80-1.70) | 3.30 (2.50-4.40) | <□0.001 | 9.018 | (6.305-12.898) |
| Sodium (mEq/L) | 136.0 (132.0-141.0) | 134.0 (130.0-140.0) | 0.045 | 0.983 | (0.966-1.000) |
| Potassium (mEq/L) | 4.10 (3.60-4.70) | 4.50 (3.70-5.30) | 0.845 | 0.997 | (0.968-1.030) |
| Bilirubin (mg/dL) | 11.54 (5.00-20.40) | 13.40 (5.80-24.00) | 0.247 | 1.008 | (0.995-1.020) |
| Aspartate Aminotransferase (AST) (IU/L) | 106.0 (66.0-175.0) | 124.0 (66.0-188.0) | 0.421 | 1.000 | (1.000-1.000) |
| Alanine Aminotransferase (ALT) (IU/L) | 44 (30-82) | 45 (31-74) | 0.088 | 0.999 | (0.998-1.000) |
| Alkaline Phosphatase (ALP) (IU/L) | 177.5 (120.0-264.0) | 181.0 (132.5-254.5) | 0.689 | 1.000 | (0.999-1.000) |
| Albumin (g/dL) | 2.50 (2.20-2.90) | 2.50 (2.10-2.80) | 0.366 | 0.913 | (0.751-1.110) |
| Globulin (g/dL) | 3.60 (2.95-4.40) | 3.30 (2.55-3.90) | <□.001 | 0.676 | (0.548-0.832) |
| 2. Female | No AKI | AKI | Univariate Logistic Regression |  |  |
|  |  |  | p-value | Odds | 95% CI of |

|  |  |  |  | ratio<br>(OR) | OR |
| --- | --- | --- | --- | --- | --- |
| Infection (Yes) | 43 (52.4%) | 19 (63.3%) | 0.306 | 1.567 | (0.663-3.701) |
| Alcohol (Yes) | 5 (6.1%) | 3 (10.0%) | 0.482 | 1.711 | (0.383-7.646) |
| ACLF Grade at admission |  |  |  |  |  |
| 1 | 9 (12.3%) | 2 (7.1%) | Reference category |  |  |
| 2 | 19 (26.0%) | 5 (17.9%) | 0.856 | 1.184 | (0.192-7.320) |
| 3 | 45 (61.6%) | 21 (75.0%) | 0.369 | 2.100 | (0.417-10.580) |
| Age (years) | 40 (30-48) | 40 (30-49) | 0.407 | 1.014 | (0.981-1.049) |
| Hemoglobin<br>(g/dL) | 8.7 (7.3-10.5) | 8.3 (6.9-9.8) | 0.549 | 0.945 | (0.786-1.140) |
| TLC (per mm<sup>3</sup>) | 9430 (5180-15201) | 12100 (6600-19740) | 0.075 | 1.000 | (1.000-1.000) |
| Platelets (x10<sup>3</sup> per<br>mm<sup>3</sup>) | 91.0 (60.0-147.0) | 82.5 (50.0-124.0) | 0.273 | 0.997 | (0.990-1.000) |
| INR | 2.700 (1.870-3.740) | 2.900 (1.730-4.080) | 0.609 | 1.057 | (0.854-1.308) |
| Urea (mg/mL) | 42.00 (27.00-64.00) | 112.00 (85.00-130.00) | <□ 0.00<br>1 | 1.024 | (1.014-1.035) |
| Creatinine<br>(mg/dL) | 1.10 (0.60-1.66) | 2.90 (2.30-3.60) | <□ 0.00<br>1 | 14.037 | (4.870-40.461) |
| Sodium (mEq/L) | 137.0 (133.0-140.0) | 134.0 (126.0-140.0) | 0.044 | 0.946 | (0.896-0.999) |
| Potassium<br>(mEq/L) | 3.80 (3.30-4.50) | 4.30 (3.70-5.40) | 0.004 | 1.780 | (1.200-2.640) |
| Bilirubin (mg/dL) | 15.70 (5.10-23.80) | 12.75 (3.10-20.10) | 0.231 | 0.975 | (0.936-1.020) |
| Aspartate<br>Aminotransferase<br>(AST) (IU/L) | 115.0 (58.0-224.0) | 116.5 (56.0-167.0) | 0.866 | 1.000 | (0.998-1.002) |
| Alanine<br>Aminotransferase | 47 (30-122) | 56 (24-98) | 0.658 | 1.001 | (0.998-1.003) |
| (ALT) (IU/L) |  |  |  |  |  |
| Alkaline Phosphatase (ALP) (IU/L) | 233.0 (133.0-337.0) | 236.0 (159.0-332.0) | 0.977 | 1.000 | (0.998-1.002) |
| Albumin (g/dL) | 2.50 (2.20-2.95) | 2.55 (2.10-2.80) | 0.458 | 0.764 | (0.374-1.560) |
| Globulin (g/dL) | 3.10 (2.55-3.80) | 3.00 (2.35-3.55) | 0.580 | 0.861 | (0.507-1.460) |

Multivariate analysis confirmed serum creatinine as the most robust independent predictor of AKI in both males (OR = 9.144) and females (OR = 13.516; *p* < 0.001 for both) (Table 3), as expected. While several other variables showed significance in univariate analysis among males, including infection, alcohol use, ACLF grade, and urea, these did not retain independent significance after adjustment, suggesting their effects may be mediated through interconnected clinical pathways.

**Table 3.** Multivariate logistic regression analysis of factors affecting presence of AKI in males and females.

| Predictor | Estimate | SE | Z | p-value | Odds ratio (OR) | 95% CI of OR |
| --- | --- | --- | --- | --- | --- | --- |
| **Males** |  |  |  |  |  |  |
| Intercept | -2.907 | 3.080 | -0.944 | 0.345 | 0.055 | (0.000-22.840) |
| Infection (Yes) | 0.132 | 0.358 | 0.368 | 0.713 | 1.141 | (0.566-2.300) |
| Alcohol (Yes) | 0.117 | 0.443 | 0.265 | 0.791 | 1.124 | (0.472-2.680) |
| **ACLF Grade at admission:** |  |  |  |  |  |  |
| 2 | -0.130 | 0.907 | -0.144 | 0.886 | 0.878 | (0.148-5.200) |
| 3 | -0.130 | 0.874 | -0.149 | 0.882 | 0.878 | (0.158-4.870) |
| Hemoglobin (g/dL) | -0.081 | 0.079 | -1.033 | 0.302 | 0.922 | (0.790-1.080) |
| Urea (mg/mL) | 0.005 | 0.005 | 1.135 | 0.256 | 1.005 | (0.996-1.010) |
| Creatinine (mg/dL) | 2.213 | 0.307 | 7.218 | <0.001 | 9.144 | (5.013-16.680) |
| Sodium (mEq/L) | -0.002 | 0.019 | -0.088 | 0.930 | 0.998 | (0.961-1.040) |
| Globulin (g/dL) | -0.346 | 0.187 | -1.852 | 0.064 | 0.707 | (0.490-1.020) |
| **Females** |  |  |  |  |  |  |
| Intercept | 0.231 | 5.921 | 0.039 | 0.969 | 1.260 | (0.000-138056.300) |
| Urea (mg/mL) | 0.002 | 0.009 | 0.199 | 0.842 | 1.002 | (0.985-1.020) |
| Creatinine (mg/dL) | 2.604 | 0.613 | 4.248 | <0.001 | 13.516 | (4.065-44.940) |
|  |  |  |  | 1 |  |  |
| Sodium (mEq/L) | -0.052 | 0.042 | -1.216 | 0.224 | 0.950 | (0.874-1.030) |
| Potassium (mEq/L) | 0.160 | 0.291 | 0.551 | 0.582 | 1.174 | (0.664-2.070) |

*OR: Odds Ratio, CI: Confidence Interval*
*Significance at the \*\*\*0.001 level, •0.1 level*

## Discussion

In our study, we observed that within an age- and ACLF-grade-matched cohort of male and female patients, kidney function parameters, specifically serum urea and creatinine, were significantly higher in males compared to females. This difference is likely attributable to a greater incidence of ACLF-associated AKI in male patients (Figure 1A-C, Table 1).

However, the severity of AKI, as assessed by AKI staging, did not differ significantly between males and females with ACLF-AKI (Figure 1D, Table 1). Similarly, the proportion of patients without AKI at admission who progressed to AKI during hospitalization was comparable across genders (Figure 1E, Table 1). Females who developed AKI exhibited a more pronounced increase in serum creatinine (δ sCr) compared to their male counterparts (Figure 1F, Table 1).

Further analysis revealed distinct gender-specific risk factors for AKI in ACLF patients. In males, serum creatinine, ACLF grade, presence of infection, alcohol use, urea, sodium, and globulin emerged as independent predictors of AKI (Table 2). In contrast, for females, serum creatinine, followed by potassium, sodium, and urea, were identified as independent predictors (Table 2). Despite these differences, multivariate analysis revealed that serum creatinine was the only consistent and independent predictor of AKI in both sexes (Table 3), as expected.

However, these findings underscored critical gender-based differences in diagnostic markers, particularly serum creatinine and δ sCr, in ACLF patients and suggested potential avenues for deeper investigation. A previous report suggests that in hospitalized cirrhotic patients with infection, females had lower baseline and peak creatinine, similar to our study, but comparable delta creatinine and were more likely to receive RRT at equivalent creatinine levels (16). Therefore, future studies should aim at elucidating the biological mechanisms underpinning these disparities, such as the modulatory roles of sex hormones like estrogen and testosterone, and the genetic influence of XX versus XY chromosomal composition on renal physiology and dysfunction. Immunological differences, including variations in cytokine profiles and immune cell activation between males and females, may also contribute to distinct responses in ACLF-AKI and warrant further exploration.

Given the steeper rise in serum creatinine (δ sCr) observed in females following AKI onset, it is important to reassess whether current diagnostic thresholds for AKI are equally valid for both genders. Gender-related differences in AKI risk and progression have been documented in other clinical contexts, with males often demonstrating higher susceptibility and worse outcomes—likely influenced by hormonal, immune, and hemodynamic factors (14,15).

Developing gender-sensitive diagnostic criteria and monitoring strategies may enhance early detection, enable timely intervention, and ultimately improve outcomes for both male and female ACLF patients.

### Limitations

A major limitation of our study was that the number of female study participants were lower than that of men. The inclusion of a larger and more balanced cohort of male and female patients would provide greater statistical power, helping to validate these findings further.

### Study approval

The study was approved by the All India Institute of Medical Sciences, New Delhi ethics committee [Approval reference no. IEC.473/07.10.2016 and IEC.369/01.08.2016/RP-20/2016] and abides by the Declaration of Helsinki. Informed and written consent was obtained from each study participant and any identifiers were withheld from the study analyses.

## Data availability

All data associated with this manuscript are available within the manuscript files.

## Acknowledgements

PA would like to extend her gratitude for the support provided by WomenLift Health through her participation in the 2023 India Leadership Journey, a yearlong leadership development experience designed for mid-to-senior career women leaders working in public and global health in India. The tools, frameworks, coaching, mentorship, and the access to a robust peer network provided by the Journey played a key role in allowing her to apply a gender lens to her research to ask new questions and obtain deeper insights.

We are grateful to the study participants and their kin for participating in this study. We are grateful to the staff and clinical personnel of the Dept. of Gastroenterology for their continued support in our research work.

## Author contributions

RS, SP: Data collection and statistical analysis, inferences and construction of figures and tables, drafting of manuscript, S.: Critical inputs into the manuscript, patient recruitment and data collection, PA: Conceptualization and study design, inferences, drafting of manuscript and overall supervision.

